# Hematologic, Immunologic and Outcome Characteristics of Severe acute respiratory syndrome 2 (SARS-COV-2) among People Living with HIV in Eastern Uganda: A Retrospective Study

**DOI:** 10.1101/2025.02.22.25322708

**Authors:** Timothy Otaala, Ronald Opito, Kenneth Mugisha, Bonniface Oryokot, Baker Bakashaba, Moses Esabu, Susan Nabadda Nbidde, Benedicto Watmon, Wilson Etolu, Moses Yovani Lubaale, Peter Olupot-Olupot

## Abstract

**Introduction:** Globally People Living with HIV (PLWHIV) are prone to severe opportunistic infections with Coronavirus disease of 2019 (COVID-19) inclusive. Dual infection of HIV and COVID-19 could complicate the clinical outcomes of these patients. This study sought to determine hematologic, immunologic, virologic and outcome characteristics of people with HIV and COVID-19 co-infections in two Mbale and Soroti Regional Referral Hospitals in Eastern Uganda.

**Methods:** A retrospective review of medical records of PLHIV and contracted COVID-19 virus was conducted in two large regional referral hospitals in Eastern Uganda using a data abstraction tool. Data was captured using Kobo collect toolbox, downloaded in Microsoft Excel and analyzed using STATA version 15.0. Descriptive statistics was reported as frequencies and proportions, while contingency and comparisons were done at the bivariate level analysis. The penalized logistic regression was conducted at multivariate level to establish the factors associated with COVID-19 severity among PLHIV.

**Results:** A total of 100 patient records had 38%(n=38) of individuals aged 40-50 years and 62%(n=62) females. Most patients (6 in 10) were peasants with 79%(n=79) having low-income levels. Other than HIV, 3 out of 10 patients had comorbidities. Also, 30 had haematological records, of which 9 (30%) had leucocytosis and 3, leucopoenia. Further, 67% had immunologic records, of which 22 (33%) had CD4 counts <200 cells/mm3. Only 22% of patients had viral load results, of which 8 (36) were unsuppressed. Nineteen percent (19/100) patients had severe COVID-19 and 14% (14/100) died. Socio-demographic factors significantly associated with severe COVID-19 outcomes were being male (P=0.026) and having other comorbidities (P<0.001).

**Conclusion:** A significant proportion of PLHIV co-infected with COVID-19 had abnormal hematological and virological status possibly due to varying socio-demographic characteristics. Clinical outcomes of HIV and COVID-19 co-infection may therefore vary depending on an interplay between host factors, viral factors, and comorbidities.

## Introduction

Sub–Saharan Africa where Uganda falls is home to 67% of the global HIV burden [1]. In Uganda, about 1.2 million people aged 15 to 64 years are living with HIV/AIDS [1]. The national HIV prevalence is 5.8% with Teso and Elgon sub regions having a prevalence of 4.2% and 3.8% respectively [2]. By 11th February 2024, 774,631,444 confirmed cases of COVID-19, including 7,031,216 deaths have been registered worldwide. In Africa, there were 9,575,413 cases and 175,495 deaths while in Uganda, there were 169,276 cases and 3,632 deaths in Uganda [3].

HIV virus attacks the body’s immune system by firstly attaching itself to the CD4 cell receptors (chemokine receptor 5 (CCR5) or chemokine receptor 4 (CXCR4). Both the direct and indirect cytopathic effects of the virus on the host’s innate immune response usually occurs during translocation from immune activation, leaking guts, immune homeostasis and immune dysregulation [4]. On the other hand, the SARS-CoV-2 virus is a β-coronavirus which mainly affects bronchial and alveolar epithelial cells, alveolar macrophages by attachment, penetration, bio synthesis, maturation and release on the Angiotensin converting enzyme 2 (ACE2) [4, 5].

People living with HIV (PLHIV) are at a risk of chronic co-morbidities, ongoing inflammation, progressive immune dysfunction, long-term infection by the virus and exposure to antiretroviral drug toxicities while COVID-19 patients are prone to cardiovascular, renal dysfunction, thrombotic, and central nervous system (CNS) complications [6]. Dual infection of the two pandemics could be catastrophic with some studies suggesting that HIV and COVID-19 co-infected patients are 21 times more likely to die of COVID-19 infection compared to persons who do not have HIV [7-9]. These patients are therefore predisposed to poor treatment outcomes and interruptions in HIV care and treatment [4]. Since March 2020, there has been growing interest and some emerging body of evidence regarding co-infections of HIV and COVID-19 [10-12]. The interactions of SARS COV-2 with other health conditions, including HIV, remain poorly studied [8, 13]. The high population of PLHIV inconclusive information of the therapeutic potential protection of HIV agents against COVID-19 and no studies that have been conducted on hematologic, immunologic, virologic characteristics and outcomes amongst HIV and COVID-19 co-infected patients in Eastern Uganda despite the poor clinical outcomes they could be inclined to, highlight a dire need to conduct this study. Understanding the Ugandan context and thereby coming up with interventions to reduce the associated morbidity and mortality amongst HIV and COVID-19 co-infected patients is significant.

This study therefore sought to determine the hematologic, immunologic, virologic and outcome characteristics of people living with HIV and COVID-19 co-infections in two large Regional Referral Hospitals in Eastern Uganda.

## Materials and Methods

### Study Design and area

This was a retrospective study which involved secondary data review of medical records of patients admitted in the COVID-19 units of the two regional referral hospitals to determine their HIV status, haematologic, immunologic, virologic profiles and clinical outcomes. Using data abstraction tool, we abstracted relevant data from the COVID-19 inpatient register, admission files and electronic databases for all the patients who had recorded HIV status and admitted in the hospital between March 2020 and Feb 2022. Data abstraction was done between June to August 2023. The study was conducted at Soroti and Mbale Regional Referral Hospitals. Soroti Regional Referral Hospital is 294km by road, northeast of Mulago National Referral Hospital, 102 km by road from Mbale Regional Referral Hospital, and 123.5km from Lira Regional Referral Hospital by road. It is a regional referral hospital for the Teso sub-region. Soroti hospital has a 250-bed capacity and was a regional COVID-19 treatment centre. The COVID-19 treatment unit had a bed capacity of 60 beds which were housed in the mental unit, Tuberculosis ward and additional tents that were evacuated to manage the big numbers. Mbale Regional Referral Hospital is located on Palisa Road, in the central business district of Mbale city. It is about 144 kilometres (89 miles), by road, northeast of Jinja Regional Referral Hospital, in the city of Jinja. Mbale hospital had a 40-bed capacity for their COVID-19 treatment unit (*www.health.go.ug*).

### Inclusion and Exclusion criteria

Patient recruitment flow chart

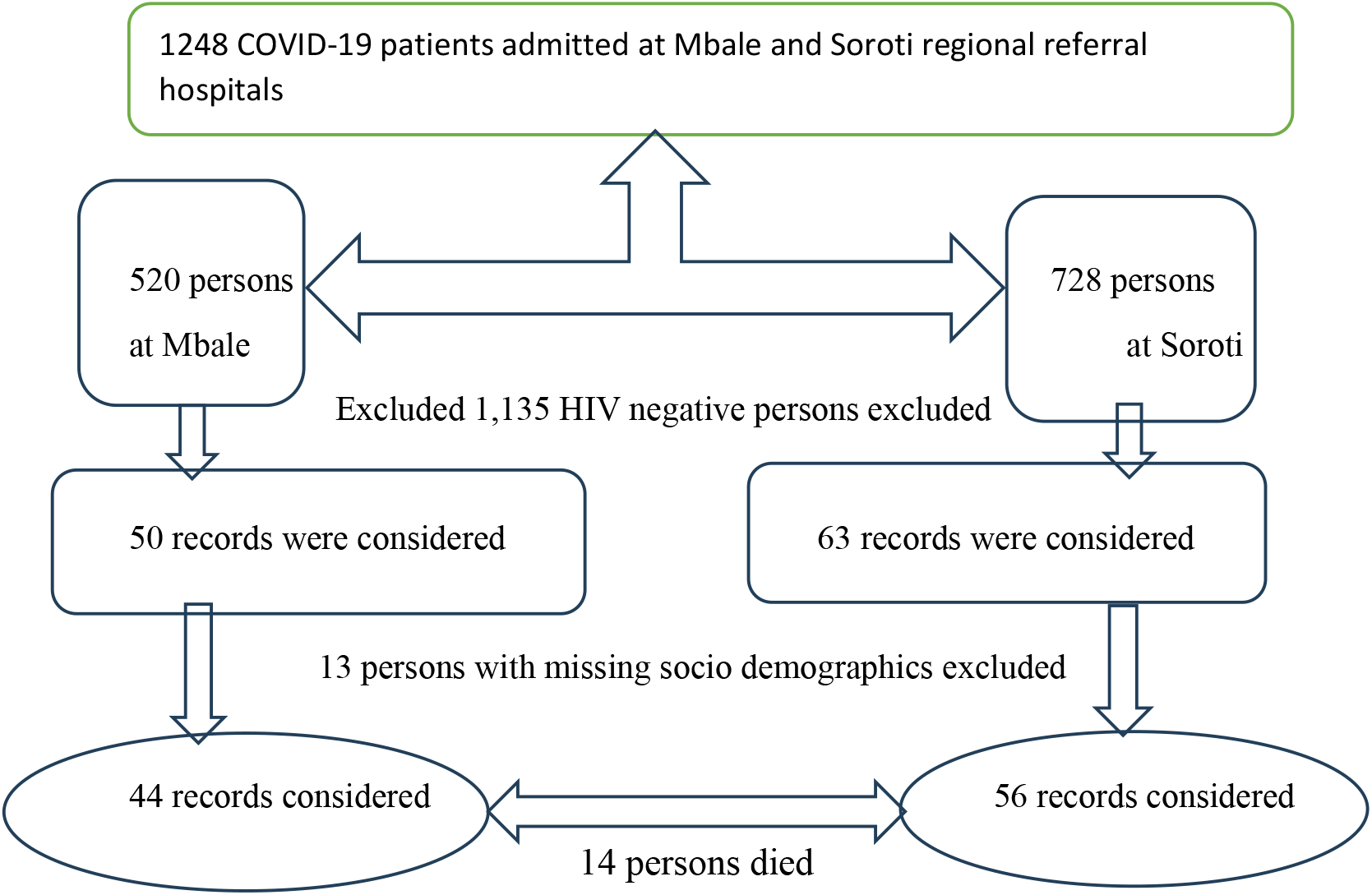

### Study Variables

The variables were extracted from the records in line with the study objectives as explained below:

Independent Variables included age, sex, marital status, religion, occupation, income status, location, their ART status, whether they had any comorbidities. The dependent variables

The primary outcome of the study was Severity of COVID-19 disease classified and measured as a binary outcome, non-severe/mild or Moderate/Severe.

The secondary outcomes included Length of Hospitalization (duration), final outcomes (recovered, died, missed appointment), Haematologic outcomes, including haemoglobin (HB), white blood cell total counts (WBC-total), Neutrophils, Lymphocytes, Monocytes, Eosinophils, Basophils, Platelets and haematological indices (Mean Cell Volume (MCV), Mean Cell Haemoglobin Concentration (MCHC), Red Blood Cells (RBC), Red Cell Width (RDW) data for HIV and COVID-19 co-infected patients.

Immunologic status; this was the recorded CD4 levels of the patients.

Virologic outcomes; this was the recorded viral load count of the admitted patients.

### Data Collection Procedures

The study involved obtaining permission to collect patient data for HIV and COVID-19 co-infected individuals from the hospital management of Mbale and Soroti hospitals following approval from the Research and Ethics Committee of Busitema University. Patient files for HIV and COVID-19 co-infected patients between 2020 and 2022 were retrieved from the records department by two health information assistants, and captured electronically using the uniform resource locator (https://ee.humanitarianresponse.info/x/pRxpohwV) that was designed using the Kobo toolbox version.

Traceability and audit of records/data to ensure internal validity was conducted by; a vertical audit of the laboratory data for traceability of Quality control data, use of retrospective patient data from equipment that underwent routine preventive maintenance and servicing and using patient data whose sample analysis was done using calibrated auxiliary equipment.

External validity in my study was addressed through: -

Including all COVID-19 patients with HIV in the study since they are few.

Following the inclusion and exclusion criteria.

All COVID-19 patients with HIV regardless of their WHO clinical staging, presence of other co-morbidities, or are severely ill but were admitted and their patient data is available was used.

### Data Management and Analysis

Data analysis was carried out at Univariate, Bivariate, and Multivariate levels. For the Univariate analysis, categorical variables were summarized using frequencies and percentages, while continuous variables were summarized using mean (standard deviation). At bivariate level, each independent variable was run against the dependent variable so as to determine the level of association using chi square or Fisher’s exact test. AT multivariable level, penalized logistic regression was applied to determine the hematologic, immunologic, virologic and outcome variables that could cause severity of COVID-19 or affect the clinical outcomes. Factors with a p-value of less than 0.2 at bivariate level analysis, were included in the multivariable analysis and 95% Confidence interval and p-values of a p-value<0.05 were as statistically significant. Data analysis was conducted using Stata version 15.0 software. Dagitty which is a browser-based environment for creating, editing, and analyzing causal diagrams (also known as directed acyclic graphs or causal Bayesian networks was used to deal with or minimize confounding factors. The model showed minimal sufficient adjustment sets containing Age for estimating the direct effect of COVID-19 patients with HIV on Alive/Dead, Admissions, COVID-19 severity, hematologic, immunologic, and virologic outcomes: Adherence to ART, Advanced HIV disease, Age, Co-morbidities, HIV drug resistance, Income status, Occupation, Sex.

## Results

### Social demographic characteristics of HIV and COVID-19 co-infected patients

Of the 100 patients in this study, 62%(n=62) were females, aged <50years, 67% (n=67). Most of the patients; 60% (n=60) were peasants with a big proportion ;79(79%) having low-income status.

More than a third of the patients (n=34/100) had comorbidities other than HIV. While only about a quarter of the patients, 26% (n=26) were vaccinated against COVID-19. A big proportion of these patients (n=77) were in WHO HIV clinical stage 1 or 2.

The sociodemographic factors significantly associated with COVID-19 severity were, Sex where the males had 3.5 odds of developing severe COVID-19 infection compared to the females (95% CI; 1.04-11.79) and presence of comorbidities, aOR=11.13 (95%CI; 2.51-49.49). The age of the participant, WHO clinical staging and COVID-19 vaccination status did not have effect on COVID-19 severity. Almost a third (29) patients had long hospital stays of more than 21 days. Fourteen percent (n=14) of patients died.

**Table 1:** Socio demographic and outcome characteristics associated with COVID-19 Severity.

| Variable | Frequency<br>(N=100) | Covid 19 Severity status |  | cOR (95% CI) | aOR (95% CI) |
| --- | --- | --- | --- | --- | --- |
|  |  | Not severe,<br>n=81 (81.0%) | Severe,<br>n=19 (19.0%) |  |  |
| Sex |  |  |  |  |  |
| Female | 62 | 55 (88.71) | 7 (11.29) | 1 | 1 |
| Male | 38 | 26 (68.42) | 12 (31.58) | 3.49 (1.26-9.65) | 3.49 (1.04-11.79) |
| **Age** |  |  |  |  |  |
| <50 years | 67 | 59 (88.06) | 8 (11.94) | 1 | 1 |
| >50 years | 33 | 22 (66.67) | 11 (33.33) | 3.58 (1.30-9.83) | 1.51 (0.45-4.99) |
| **Occupation** |  |  |  |  |  |
| Formal | 17 | 14 (82.35) | 3 (17.65) | 1 | 1 |
| Non-formal | 83 | 67 (80.72) | 16 (19.28) | 1.01 (0.28-3.66) | 2.02 (0.37-10.84) |
| **Religion** |  |  |  |  |  |
| Christian | 86 | 69 (80.23) | 17 (19.77) | 1 |  |
| Non-Christian | 14 | 12 (85.71) | 2 (14.29) | 0.79 (0.19-3.41) |  |
| **Income status** |  |  |  |  |  |
| Low | 79 | 65 (82.28) | 14 (17.72) | 1 | 1 |
| Middle | 21 | 16 (76.19) | 5 (23.81) | 1.51 (0.49-4.62) | 2.14 (0.49-9.25) |
| **Comorbidities present** |  |  |  |  |  |
| No | 66 | 62 (93.94) | 4 (6.06) | 1 | 1 |
| Yes | 34 | 19 (55.88) | 15 (44.12) | **11.04 (3.44-35.43)** | **11.13 (2.51-49.49)** |
| **WHO Stage** |  |  |  |  |  |
| Early (1,2) | 77 | 66 (85.71) | 11 (14.29) | 1 | 1 |
| Advanced (3,4) | 23 | 15 (65.22) | 8 (34.78) | 3.17 (1.11-9.01) | 0.77 (0.16-3.69) |
| **COVID19 vaccination** |  |  |  |  |  |
| Not vaccinated | 74 | 57 (77.03) | 17 (22.97) | 1 | 1 |
| Vaccinated | 26 | 24 (92.31) | 2 (7.69) | 0.34 (0.08-1.37) | 0.86 (0.17-4.44) |
| **Length of stay** |  |  |  |  |  |
| Long (21 days and more) | 29 | 20 (68.97) | 9 (31.03) |  |  |
| Short/moderate (<20 days) | 71 | 61 (85.92) | 10 (14.08) |  |  |
| **Outcomes** |  |  |  |  |  |
| Died | 14 | 5 (35.71) | 9 (64.29) |  |  |
| Alive | 86 | 76 (88.37) | 10 (11.63) |  |  |

### Hematologic, immunologic and Virologic characteristics of HIV and COVID-19 co-infected patients

Out of 100 patients in the study, 30 patients had complete blood count results. Of these, 9 had elevated white blood cell (WBC) counts while 21 had normal/low WBC counts. 20 out of 100 patients had platelet counts. Of these, 5 patients had elevated platelet counts while 25 had low/normal counts. 25 out of 100 patients had neutrophil counts. Of these, 11 had neutrophilia (elevated neutrophil counts) while 14 had low/normal neutrophil counts. 25 out of 100 patients had lymphocyte counts. Amongst these, 24 out of 30 of the patients had normal/low lymphocyte counts. Out of 100 patients, 67 patients had CD4 counts. Of these, 22 had CD4 counts that were <200 cells/mm3 while 45 had CD4 counts that were >200. Virologic laboratory characteristics of HIV and COVID-19 co-infected patient. 22 out of 100 patients had viral load counts. Of these, 8 out of 22 had viral load counts that were >1000 copies/ul (non-suppressed) while 14 had viral load counts that were <1000 copies/ul (suppressed).

**Table 2:**
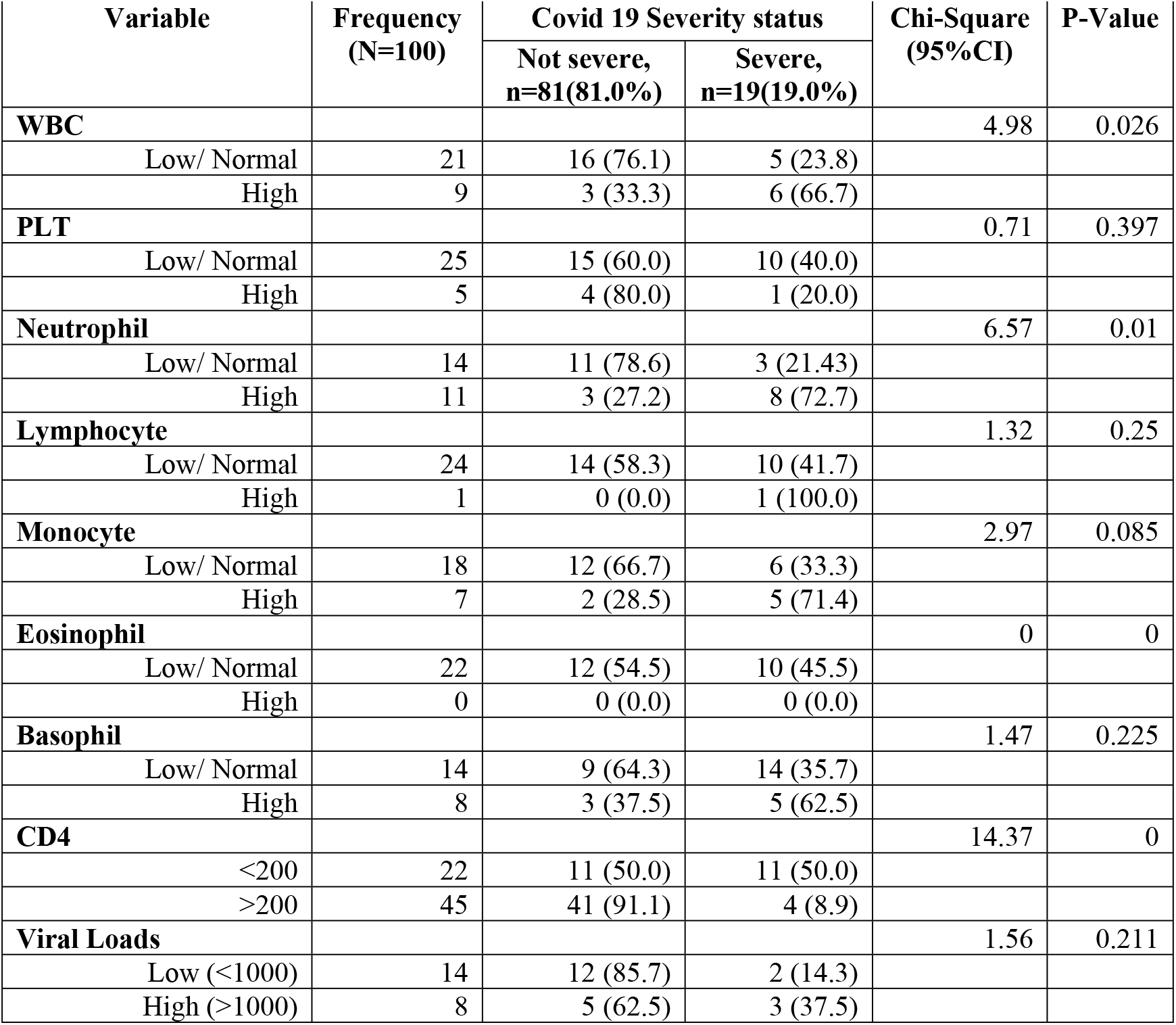
Hematologic, immunologic and Virologic characteristics of HIV and COVID-19 co-infected patients.

## Discussion

Our findings revealed significant sociodemographic disparities among HIV and COVID-19 co-infected patients, with females (62%) and individuals aged<50 years being predominant. Most patients (60%) were peasants with low-income status. These sociodemographic factors not only influenced the severity of COVID-19 but also underscored the pre-existing vulnerabilities.

Our study found a significant association between the presence of comorbidities and the severity of COVID-19. These findings align with systematic reviews by [14], which emphasize the role of comorbidities in worsening health outcomes. The contrast between comorbidities in our cohort (tuberculosis, cryptococcal meningitis) and those in developed countries (cardiovascular diseases) further illustrates the need for context-specific interventions [15, 16].

In this study, hematologic changes, such as leukocytosis (30%), neutrophilia (40%), and lymphocytopenia (46.7%) were likely exacerbated by the combined effects of HIV and COVID-19, both of which are known to affect immune function. The hematologic changes we observed underscore the importance of continuous monitoring of blood cell counts in HIV and COVID-19 co-infected patients. Co-infection may amplify pre-existing conditions such as thrombocytopenia and anaemia, commonly seen in HIV-positive individuals. These abnormalities could potentially worsen due to the added immune suppression caused by COVID-19 [17, 18]. The findings suggest that timely intervention is critical for minimizing morbidity and mortality in this high-risk population.

Furthermore, our findings highlight the compromised immune status of co-infected patients, with 22 out of 67 patients having CD4 counts below 200 cells/mm^3^, indicating advanced immunosuppression. While HIV alone is known to deplete CD4 cells, co-infection with COVID-19 appears to accelerate this depletion, possibly leading to more severe clinical outcomes. These observations align with research by Copertino, Casado Lima [19], who noted the potential for worsened immune function in co-infected individuals. The interaction between HIV and COVID-19 may trigger an exaggerated inflammatory response or “cytokine storm,” which could explain the increased morbidity and mortality seen in some patients. Our study did not specifically measure inflammatory markers like cytokines, but future research could focus on these variables to better understand the immunopathology of co-infection.

Among our cohort, 36% of patients had unsuppressed HIV viral loads (>1000 copies/µL), which could have contributed to their heightened susceptibility to severe COVID-19. HIV viral suppression is essential for maintaining immune function, and co-infection with COVID-19 may disrupt viral replication dynamics, leading to poorer clinical outcomes. The variation in viral load suppression rates between our study and that of Asghar, Haider Kazmi [20] in Nairobi (14% unsuppressed vs. 36% in our study) can be attributed to socioeconomic disparities, such as access to healthcare and adherence to antiretroviral therapy (ART). Understanding how HIV and SARS-CoV-2 interact within co-infected individuals remains an area requiring further investigation, particularly regarding how this interaction might drive viral evolution and mutation rates. The clinical outcomes in our study revealed that 14% of patients died, while 19% experienced severe COVID-19. Prolonged hospital stays were observed in nearly half of the patients, indicating that co-infection leads to more complicated disease courses. Our mortality rate aligns with findings by Ssentongo, Heilbrunn [21], who reported a 12.65% mortality rate in co-infected patients. The severe clinical outcomes in our study population were exacerbated by underlying HIV clinical staging 3 and 4 and co-infections such as Tuberculosis, Cryptococcal Meningitis and Non communicable diseases. This highlights the urgent need for targeted public health interventions to improve outcomes for this vulnerable population.

## Conclusion

Our findings demonstrated that most of the PLHIV presenting with HIV and COVID-19 co-infection were females, peasants with low-income status. Neutrophilia, Leukocytopenia, reduced CD4 counts, increased viral load cells counts were observed with 14 out of 100 of these patients dying and 19 and out of 100 developing severe COVID-19. COVID-19 severity was associated with Sex, Income status, Presence of other comorbidities, long hospital duration, and WHO clinical staging. Most HIV Positive patients with Leucocytosis, Neutrophilia, and Lymphopenia will develop severe COVID-19. Diabetes, Tuberculosis, Cryptococcal Meningitis, and Hypertension accounted for most of the co-morbidities.

This paper highlights the importance of context and vulnerabilities. We see socio demographic factors such as income, having a significant impact in an African setting. The contrast between comorbidities in our cohort (tuberculosis, cryptococcal meningitis) and those in developed countries (cardiovascular diseases) further illustrates the need for context-specific interventions. The hematologic changes that were observed in our study, underscore the importance of continuous monitoring of blood cell counts in HIV and COVID-19 co-infected patients. Timely interventions are therefore critical to abate morbidity and mortality in this high-risk population. There is therefore a dire need for HIV service providers to routinely screen, identify, and treat PLHIV for comorbidities in order to avoid the development of severe disease outcomes such COVID-19 disease and others. Further studies could focus on understanding the immunopathology of co-infection especially that the interaction between HIV and COVID-19 could have triggered an exaggerated inflammatory response or “cytokine storm,” which could explain the increased morbidity and mortality seen in some patients.

## Data Availability

All data produced are contained in the manuscript

## Author contributions

### Conceptualized and wrote the original draft

Otaala

### Conceived and supervised the overall study

Provided expert guidance and critically revised the manuscript for important intellectual content: Professor Peter Olupot-Olupot

### Data collection process

Dr Wilson Etolu.

### Manuscript writing

Dr Susan Nabadda Nbidde, Dr Mugisha Kenneth, Dr Watmon Benedicto and Dr Bakashaba Baker

### Statistical analysis, contributed to interpretation of the data, provided guidance on the study design and critically reviewed the manuscript

Professor Yovani Lubaale Moses, Dr Opito Ronald and Moses Esabu.

All authors contributed to editing the paper and approved the final submission.

## Acknowledgements

We do acknowledge the support from the management and health care professionals from Mbale and Soroti regional referral hospitals for their various contributions to this work, especially Ambrose Kiberu, Tonny Brian Makoko, Silas Okullo and Dr Mutoo Paul Bukhota.

## Study limitations

Our data were collected from a single geographic region in eastern Uganda, which may limit the generalizability of our findings to other populations. Missing variables, such as laboratory data on inflammatory markers, further constrained our analysis. Future research should consider expanding the scope to include these markers and examine the role of pre-existing hematologic conditions.

## Availability of data and materials

The study is available by request to the corresponding author

## Compliance with ethical standards

The study conformed to provisions of ethical standards in Uganda.

## Ethical approval and consent to participate in the study

Ethical consideration was obtained after clearance from Busitema University higher degrees research council. The study was approved by the Mbale Regional Referral Hospital Research and Ethics Committee. Administrative clearance was obtained from the respective hospital heads. Confidentiality and privacy of patient information was conducted throughout the study and after the study by using unique identifiers, using computers with pass words and keeping the information under lock and key.

## Consent to Publish

The Mbale Clinical Research Institute (MCRI, www.mcri.ac.ug), a research entity affiliated to the Uganda National Health Research Organization, approved publication of the manuscript.

## Abbreviations

**Term Definition**

ACE2: Angiotensin-converting enzyme 2
AHD: Advanced HIV disease
ART: Anti-retroviral therapy
CD4: Cluster of Differentiation four
COVID-19: Coronavirus 2019 Disease
CXCR4: Chemokine receptor 4
HAART: Highly Active Antiretroviral therapy
HB: Haemoglobin
HIV: Human Immune Virus
ICU: Intensive Care Unit
MCHC: Mean Cell Haemoglobin Concentration
MCV: Mean Cell Volume
PEPFAR: The U.S. President’s Emergency Plan for AIDS Relief
PLWH: Persons living with HIV
RBC: Red Blood Cells
RDW: Red Cell Width
SARS-CoV-2: Severe acute respiratory syndrome coronavirus 2
SRRH: Soroti Regional Referral Hospital
UNAIDS: United Nations Program on HIV/AIDS
UPHIA: Uganda Population Based-HIV-Impact-Assessment
WBC-total: White blood cell total counts
WHO: World Health Organization

